# Policy gradient-guided ensemble learning for enhanced polygenic risk prediction in ultra-high-dimensional genomics

**DOI:** 10.1101/2025.09.23.25336425

**Authors:** Lihang Ye, Nan Lin, Ying Luo, Liubin Zhang, Meng Liu, Wenjie Peng, Xuegao Yu, Miaoxin Li

## Abstract

Polygenic diseases challenge genetic risk prediction due to extreme dimensionality, low per-variant effect sizes, and non-additive interactions. Conventional marginal *P*-value-based methods potentially overlook subtle signals and complex dependencies, while inefficient random sampling in ensembles misses sparse signals. We introduce ELAG, an ensemble learning framework that advances feature bagging by reformulating variant selection as an approximate reinforcement learning problem. Leveraging policy gradients, a parameterized policy optimizes adaptive sampling—evolving beyond uniform strategies like random forests through importance-distribution guidance—to direct non-linear classifiers toward synergistic variants. This enables scalable navigation of millions of loci without pre-filtering, handling intricate architectures. In high-polygenicity, low-heritability simulations, ELAG boosted predictive accuracy (ΔAUROC = 0.0644). For neuro-immune diseases like rheumatoid arthritis, it enhanced AUROC from 0.6866 to 0.7354 and polygenic scores (e.g., Lassosum AUROC 0.7186 → 0.7543), outperforming mainstream methods. ELAG is robust to missing data, integrable with covariates, and yields interpretable variant sets enriched for pathways and networks. It can replace random sampling with learned guidance, advancing machine learning for ultra-high-dimensional data such as genetic risk prediction.

## Main

Accurate prediction of complex polygenic diseases holds immense potential for preventive medicine, particularly for chronic conditions like Alzheimer’s disease, rheumatoid arthritis, and multiple sclerosis, where early intervention can fundamentally alter disease trajectories^1-6^. However, translating this potential into clinical reality is hindered by two fundamental and conflicting challenges inherent to genomic data. First, its extreme dimensionality—often millions of variants versus thousands of individuals—creates a severe *p* >> *n* problem that elevates the risk of overfitting^7,8^. Second, the genetic architecture of these diseases is profoundly complex, driven by myriad variants with small, non-additive effects that defy simple linear models^9^. The central task of genomic prediction is therefore to navigate the tension between aggressively reducing dimensionality and faithfully preserving this complex biological signal.

To manage dimensionality, virtually all prediction pipelines, from polygenic risk scores (PRS)^10-12^ to machine learning (ML) classifiers^13-18^, always rely on a preliminary feature filtering step. The de facto standard is the *P*-value thresholding approach (PTA) from genome-wide association studies (GWAS)^19-22^. While computationally tractable, PTA directly collides with the challenge of genetic complexity. By operating on marginal, additive statistics, it imposes a coarse, linear filter on a non-linear system, systematically overlooking variants whose importance emerges through interaction and is susceptible to selection bias (“winner’s curse”)^23^.

Attempts to move beyond PTA have consistently failed to resolve the field’s central tension. Wrapper methods, such as ant-colony (ACO)^24^ or genetic algorithms (GA)^25^, promise a more direct optimization of predictive performance but become computationally intractable at the genomic scale, forcing a reliance on the very PTA step they seek to replace. Integrated methods like decision-tree-based random forests^26,27^ can capture non-linearities but face their scalability ceiling. In the ultra-high-dimensional genomic setting, their intrinsic random subspace sampling becomes inefficient, diluting the signal from causal variants to the point where an upstream PTA step is again required for feasible application^28,29^. This creates a pervasive methodological bottleneck: the field remains dependent on a filtering strategy that is fundamentally misaligned with the complex architectures we aim to model.

Overcoming this bottleneck highlights the need for methodological advances. From the field of reinforcement learning, policy-gradient (PG) methods offer a powerful framework for optimizing complex decision-making strategies, from large language models^30^ to experimental design in genomics^31-33^. Instead of static rules, PG learns a ‘policy’ that is iteratively refined to maximize a cumulative reward. Separately, in genomic machine learning, bootstrap aggregation (bagging) is a well-established ensemble technique for improving robustness and generalization in the face of high dimensionality^34-36^. However, a key challenge remains that standard bagging relies on random feature sampling or simple aggregation, which is inefficient for refined signal detection. Incorporating PG principles to optimize bagging represents a promising approach, potentially overcoming the bottleneck of marginal *P*-value filtering and enhancing feature selection, robustness, and generalization.

Here, we introduce ELAG (Ensemble Learning with Adaptive Sampling Guided by Policy Gradient), a framework that operationalizes this concept. We demonstrate through extensive simulations that ELAG outperforms the baseline, particularly in challenging scenarios of high polygenicity and small sample sizes. We then validate ELAG on three neuro-immune disorders (Alzheimer’s disease, rheumatoid arthritis, and multiple sclerosis) in the UK Biobank (UKBB)^37^, showing that it consistently improves prediction performance over mainstream methods. We also demonstrate its robustness to genotype missingness and the ability to incorporate non-genetic covariates. Finally, ELAG produces biologically interpretable variant sets and enhances conventional PRS models, providing a versatile and clinically applicable tool for genomic risk prediction.

## Results

### Overview of ELAG

ELAG is a machine learning framework designed to overcome the limitations of static feature selection in genomic risk prediction (Figure 1). ELAG operationalizes a dynamic, learning-based approach by using PG optimization to guide adaptive sampling within an ensemble of bagged classifiers. This design enables the adaptive enrichment of informative variants from a vast pool, maintaining computational tractability while enhancing predictive power, particularly in challenging high-polygenicity and small-sample-size scenarios.

**Figure 1.**
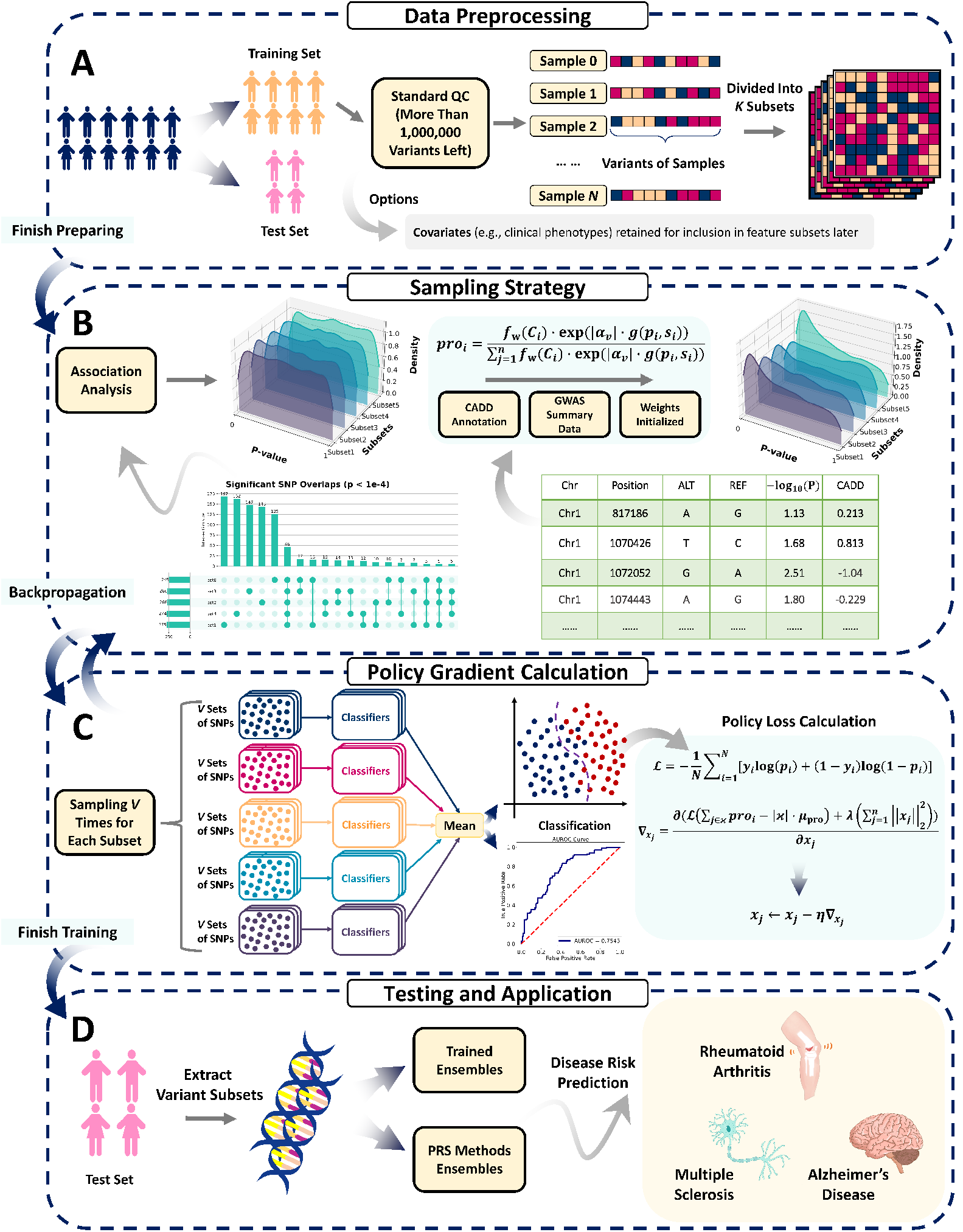
Overview of the ELAG framework. The workflow of ELAG consists of four main stages. **A**. Data Preprocessing. The full cohort is split into training and test sets. After standard quality control (QC), the training data is divided into *K* subsets for cross-validation. Though ELAG is designed for genomic data initially, it also accepts covariates. Covariates can be concatenated with the variant subsets sampled by ELAG, and the combined feature vectors can be used for classifier training and evaluation. **B**. Sampling Strategy. Initially, a policy, initialized with information (e.g., GWAS *P*-values, CADD scores), defines a sampling probability for every variant. **C**. Policy Gradient Calculation. Multiple variant subsets are drawn from probability distribution sets. Ensemble classifiers are trained on each subset, and their aggregated predictive loss is calculated. Policy loss is computed to update the policy’s parameters, refining the sampling probabilities to favor variants improving model performance. The loop continues until training is completed. **D**. Testing and Application. The final trained ensembles are applied to the independent test set for disease risk prediction. The optimized variant subsets discovered by ELAG can also be extracted to enhance the performance of PRS methods. Thus, the framework is modular, allowing for flexible choices of input priors and downstream predictors.

The core mechanism of ELAG replaces the *P*-value thresholding of conventional pipelines with a learnable, probabilistic sampling policy. This policy assigns a sampling probability to each variant, initialized using GWAS summary statistics and optionally augmented with other biological priors (e.g., CADD scores^38^). During training within each bag, diverse variant subsets are sampled according to the current policy. These subsets are used to train base classifiers (e.g., XGBoost) capable of capturing complex, non-linear genotype-phenotype relationships. The predictive performance of the classifier ensemble serves as a reward signal, which is then used to update the policy parameters via a PG-inspired algorithm. This creates a synergistic loop: the policy is iteratively refined to favor variants that contribute to better prediction, while the bagging procedure ensures the reward signal is robust and promotes a generalizable solution.

The final output of ELAG is a trained ensemble of models ready for individual risk prediction on independent test data. Furthermore, ELAG can function as a modular feature-selection engine. The prediction-optimized variant sets it identifies can be extracted and integrated into established downstream tools, such as conventional PRS methods, to significantly boost their performance. A detailed description of the methodology is provided in the Methods section.

### Simulation studies demonstrate ELAG’s superior predictive performance

We performed extensive simulations to systematically benchmark ELAG against a standard PTA (as baseline)^39^. We designed scenarios to probe the method’s performance across key challenges in genetic prediction: varying heritability, sample size, and the degree of polygenicity (number of causal QTLs), while also assessing its compatibility with different classifiers and its ability to leverage external information (Figure 2), based on a linear–nonlinear mixed model (LNMM). Unless specified otherwise, simulations used a balanced case-control design, pairwise in low linkage disequilibrium (*r*^2^ < 0.1), and XGBoost as the base classifier. More simulation settings are described in Methods.

**Figure 2.**
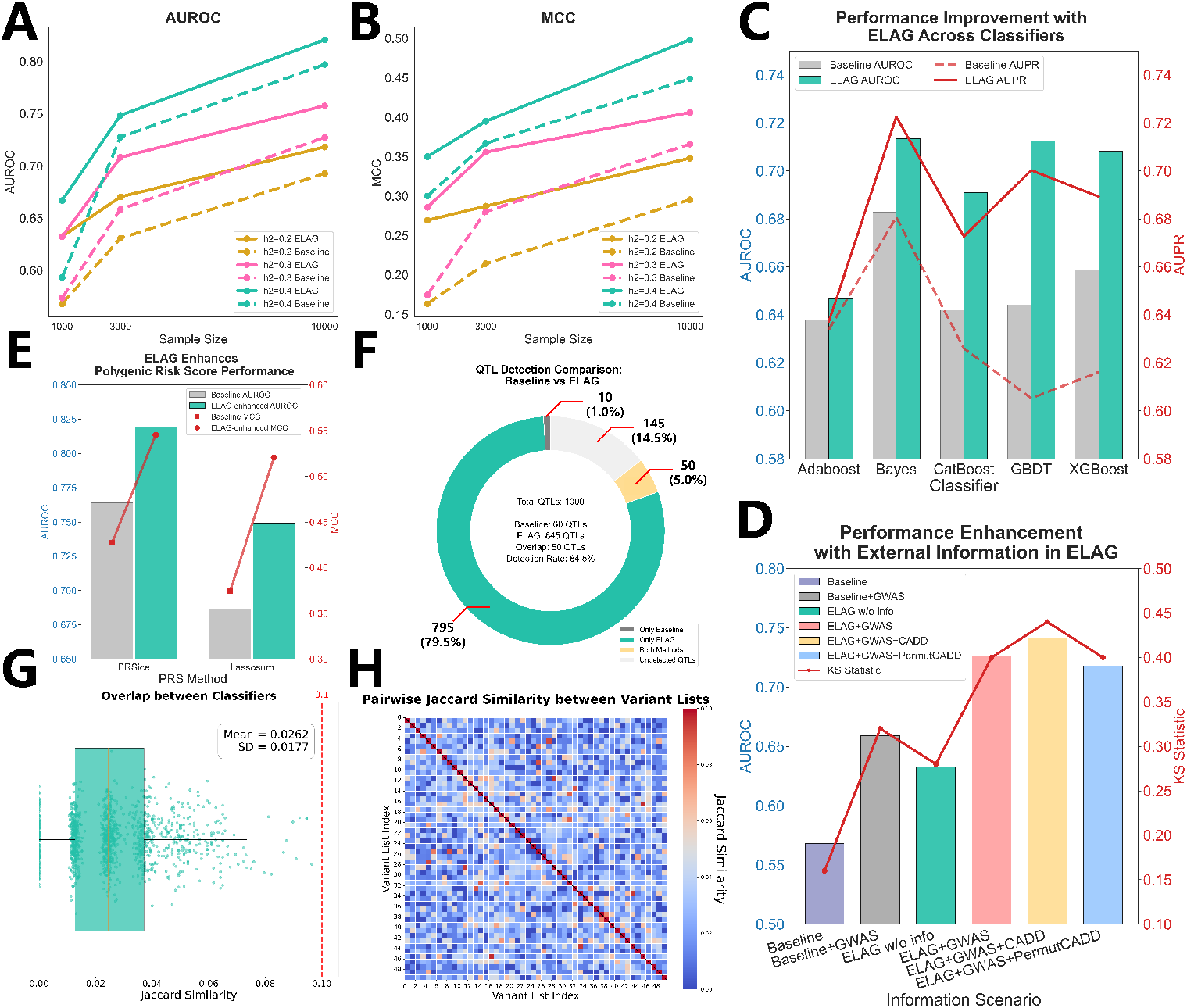
Results of simulations. **A, B**. Comparison of AUROC and MCC between the baseline method (dashed lines) and ELAG (solid lines) across varying sample sizes and heritability (*h*^2^) levels. ELAG’s advantage is most pronounced in low-power settings (small sample size, low *h*^2^). **C**. Performance comparison (AUROC and AUPR) between ELAG combined with various machine learning classifiers and the baseline method, demonstrating ELAG’s model-agnostic versatility. **D**. Performance gains (AUROC and KS statistic) from integrating external priors. External GWAS and functional scores improve ELAG’s predictive accuracy, with robustness preserved under noisy annotations. **E**. Improvement in predictive performance (AUROC and MCC) of two standard PRS methods (PRSice, Lassosum) when using variant subsets selected by ELAG. **F**. Comparison of true causal QTLs identified by the baseline method versus the collective set identified across all classifiers in the ELAG ensemble. ELAG recovers a vastly larger number of true signals. **G, H**. Distribution and heatmap of pairwise Jaccard similarity for the QTL sets selected by classifiers in ELAG. The low similarity indicates that ELAG achieves its high QTL recovery rate by selecting diverse and complementary feature sets.

### ELAG excels in low-power and highly polygenic scenarios

A primary challenge in genomics is detecting subtle genetic signals in cohorts of limited size. We first evaluated ELAG in such settings, simulating traits with 1,000 causal QTLs under varying sample sizes. As demonstrated across different heritability levels (Figure 2A and 2B and Supplementary Table 1), ELAG’s performance advantage over the PTA baseline was most pronounced at smaller sample sizes. For a sample size of 1,000, ELAG delivered substantial gains across all heritability levels (e.g., ΔAUROC = 0.0588–0.0736). The improvement was particularly striking for the low heritability (*h*^2^=0.2) trait, where ELAG achieved a 64.9% relative improvement in the Matthews Correlation Coefficient (MCC). The difference attenuated with larger sample sizes, with AUROC improvements reduced to 0.0237–0.0305 at the sample size of 10,000.

**Table 1.** Performance comparison of ELAG, PTA, ACO and GA across three diseases. Multi-metric classification performance under the best model configurations is reported, highlighting ELAG’s superior power and robustness across AD, RA and MS cohorts.

| Disease | Method | AUROC | AUPR | KS | ACC | MCC | Precision | Recall | F1-Score |
| --- | --- | --- | --- | --- | --- | --- | --- | --- | --- |
| AD | **ELAG** | **0.7237** | **0.7421** | **0.3521** | **0.6714** | **0.351** | **0.641** | **0.7793** | **0.7034** |
|  | PTA | 0.6841 | 0.6942 | 0.3099 | 0.6502 | 0.3 | 0.6539 | 0.6385 | 0.6461 |
|  | ACO | 0.6699 | 0.6868 | 0.2582 | 0.6268 | 0.2542 | 0.6364 | 0.5915 | 0.6131 |
|  | GA | 0.6808 | 0.6898 | 0.2911 | 0.6455 | 0.2972 | 0.6824 | 0.5446 | 0.6057 |
| RA | **ELAG** | **0.7354** | **0.7284** | **0.3663** | **0.6832** | **0.3813** | **0.6434** | **0.8218** | **0.7217** |
|  | PTA | 0.6866 | 0.6876 | 0.3267 | 0.6584 | 0.327 | 0.627 | 0.7822 | 0.696 |
|  | ACO | 0.6542 | 0.6664 | 0.2475 | 0.6238 | 0.3023 | 0.7907 | 0.3366 | 0.4722 |
|  | GA | 0.6241 | 0.6226 | 0.2574 | 0.6287 | 0.2817 | 0.7167 | 0.4257 | 0.5342 |
| MS | **ELAG** | **0.7185** | **0.6953** | **0.3537** | **0.6768** | **0.3533** | **0.6611** | **0.7256** | **0.6919** |
|  | PTA | 0.6556 | 0.6287 | 0.2927 | 0.6433 | 0.287 | 0.6358 | 0.6707 | 0.6528 |
|  | ACO | 0.6511 | 0.6518 | 0.2378 | 0.6189 | 0.2382 | 0.6127 | 0.6463 | 0.6291 |
|  | GA | 0.6478 | 0.6154 | 0.2622 | 0.6311 | 0.2628 | 0.6229 | 0.6646 | 0.6431 |

The advantage of ELAG was also magnified in highly polygenic architectures, where individual variant effects are weaker (Supplementary Table 2). With 1,000 QTLs and a sample size of 1,000, ELAG improved AUROC by 0.064 over PTA. In contrast, for a less polygenic trait with only 100 QTLs, the gain was a modest 0.011. As expected, these results may highlight a fundamental limitation of PTA: its reliance on strong marginal effects causes it to fail when signals are subtle and distributed across many loci. ELAG’s adaptive sampling, however, may effectively enrich for these weaker, cumulative signals, conferring a distinct advantage in the very scenarios that are most challenging for genetic discovery.

**Table 2.**
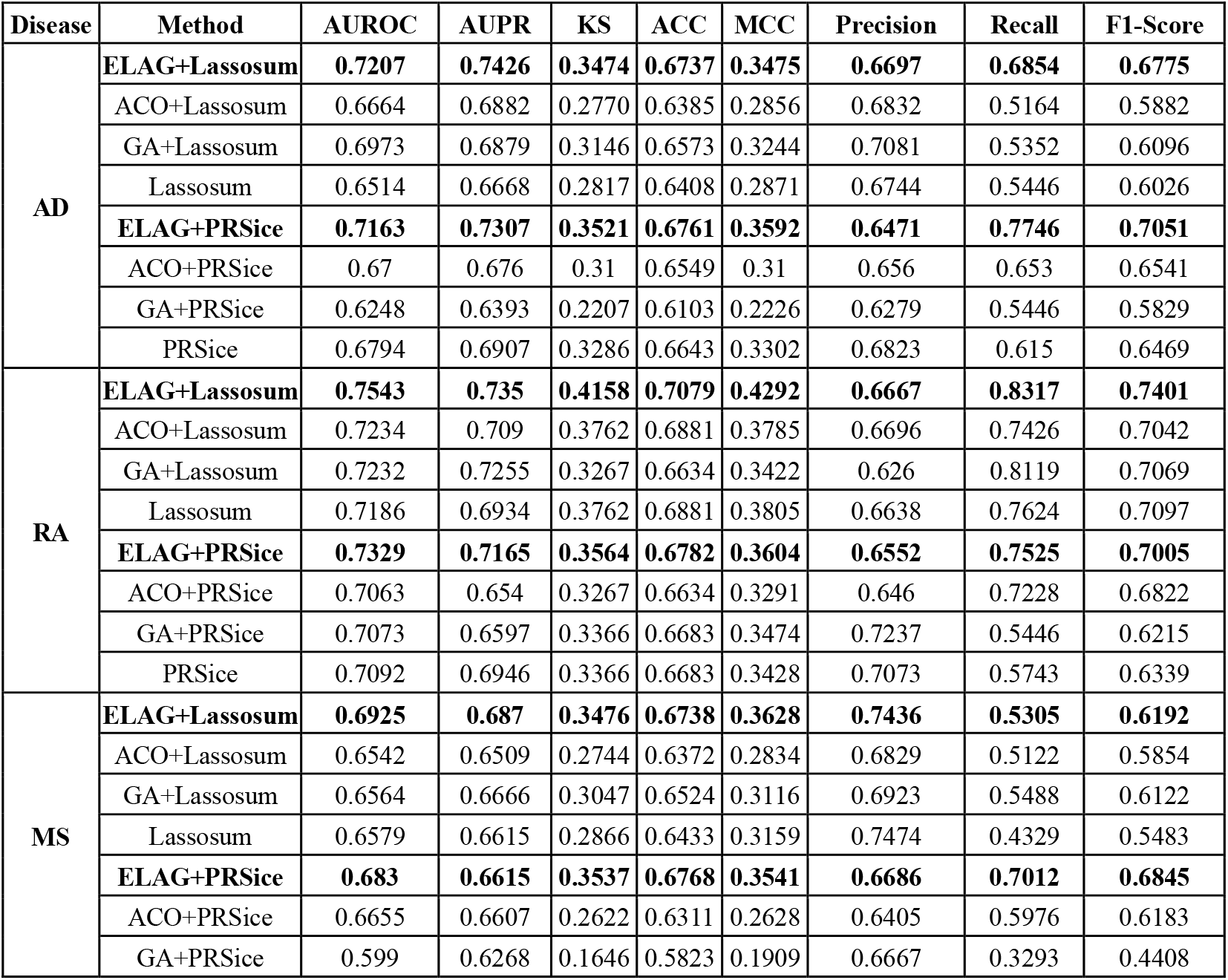
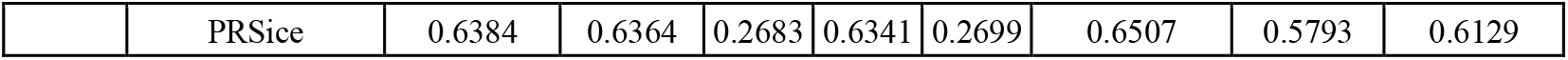
Performance metrics for three diseases (AD, RA, MS) using feature-selected PRS (feature selection method + PRS) versus standard PRS. ELAG-integrated PRS models outperform both standalone PRS and other wrapper-based feature selection approaches across AD, RA and MS, demonstrating enhanced discriminative power and robustness in polygenic risk prediction.

### Framework versatility and integration of external priors

We next confirmed that ELAG’s benefits are not specific to a single classifier. We tested its performance as an enhancement module for five different ML models, including AdaBoost, Naïve Bayes, CatBoost, and Gradient Boosting Decision Tree (GBDT). In a moderately powered setting (*N*=3,000), ELAG consistently and substantially boosted the predictive performance of every classifier tested (Figure 2C and Supplementary Table 3). For instance, it increased the AUROC of XGBoost from 0.6585 to 0.7082 and CatBoost from 0.642 to 0.691, potentially demonstrating its utility as a versatile, model-agnostic front-end. More details can be seen in Supplementary Note 1.1.

**Table 3.** Predictive performance of classifiers retrained using ELAG-aggregated top 1% variants. For each disease (AD, RA, MS), classifiers were retrained on the training set using only variants in the top 1% of aggregated XGBoost importance gains (zero-gain variants excluded) and evaluated on independent held-out test sets.

| Disease | AUROC | AUPR | KS | ACC | MCC | Precision | Recall | F1-Score |
| --- | --- | --- | --- | --- | --- | --- | --- | --- |
| AD | 0.6683 | 0.6694 | 0.277 | 0.6385 | 0.2772 | 0.6335 | 0.6573 | 0.6452 |
| RA | 0.7088 | 0.6686 | 0.3465 | 0.6733 | 0.3551 | 0.6423 | 0.7822 | 0.7054 |
| MS | 0.6789 | 0.6642 | 0.2866 | 0.6433 | 0.2866 | 0.6424 | 0.6463 | 0.6444 |

Beyond its adaptability, ELAG can seamlessly integrate external knowledge to guide its sampling policy. To test this, we used a low-power scenario (*N*=1,000, *h*^2^=0.2) where priors from a larger GWAS (*N*=40,000) and informative functional annotations (CADD-like scores) were available. Incorporating GWAS statistics alone dramatically improved ELAG’s AUROC from 0.6324 to 0.7264 (Figure 2D and Supplementary Table 4). The addition of informative functional scores provided a further boost to an AUROC of 0.7412. As a critical negative control, introducing shuffled scores reduced performance as expected (AUROC = 0.7184, KS = 0.40), yet ELAG still outperformed PTA supplemented with GWAS information (AUROC = 0.6592, KS = 0.32). These results demonstrate that ELAG may effectively harness external resources to further refine its search for causal variants to enhance predictive performance, while maintaining robustness in the presence of imperfect or misleading functional annotations.

### ELAG enhances standard polygenic risk score methods

To assess its translational potential, we evaluated ELAG as a feature engineering front-end for two widely used PRS methods, PRSice^40^ and Lassosum^41^. We used the variant subsets enriched by ELAG to construct ensemble PRS models. This strategy yielded dramatic performance improvements over the standard application of these tools (Figure 2E and Supplementary Table 5). For PRSice, ELAG-selected variants increased the AUROC from 0.7640 to 0.8192 and the MCC from 0.4270 to 0.5454 in a local cohort of 500/500 cases/controls and an external GWAS sample of 20,000/20,000. Lassosum, which had a lower baseline, saw its MCC climb from 0.3746 to 0.5204. These results show that by replacing the coarse, PTA-based filtering inherent in standard PRS workflows with ELAG’s adaptive selection, we may unlock a more diverse and potent set of predictive variants, significantly enhancing the utility of these established tools.

### Diverse feature selection underlies ELAG’s performance gain

Next, we investigated the mechanism behind ELAG’s superior performance. In a challenging simulation (*h*^2^=0.2, 1,000 QTLs, *N*=1,000), we compared the variants selected by an optimized PTA pipeline against those selected across ELAG’s ensemble. The PTA using the top 100 variants identified a total of 60 true QTLs. In stark contrast, the 50 base classifiers with 100 feature-size sampling in ELAG collectively identified 845 distinct QTLs (Figure 2F). This vast improvement in coverage was achieved through profound diversity; the QTLs chosen by any two classifiers in the ensemble were almost entirely different, with a mean Jaccard similarity of just 0.0262 (Figure 2G and 2H). This suggests that while PTA may converge on a small set of variants with the strongest marginal signals, ELAG’s adaptive, ensemble-based search may encourage the exploration of a much wider range of weaker, complementary signals. This diversity in feature selection may be the key mechanism enabling ELAG to capture a more comprehensive picture of a trait’s complex genetic architecture.

### Ablation Study

Finally, we conducted an ablation study (*h*^2^=0.2, 1,000 QTLs) to analyze the contribution of different modules in ELAG. Specifically, we introduced a variant without the policy gradient (VWPG), which relies only on bagging and sampling, and compared it against both the simple *P*-value filtering baseline and the full ELAG model (Supplementary Table 6). With a sample size of *N*=1000, ELAG achieved an AUC-ROC of 0.6324, compared to 0.6008 for MWPG and 0.5680 for the baseline. Similarly, the KS statistic improved from 0.16 (baseline) and 0.20 (MWPG) to 0.28 with ELAG, while MCC increased from 0.1633 (baseline) to 0.2209 (MWPG) and 0.2693 (ELAG). As expected, at larger sample sizes, the superiority of ELAG declined. For instance, at *NN*=10000, ELAG reached an AUC-ROC of 0.7180 and an MCC of 0.3481, outperforming MWPG (0.7123/0.3220) and the baseline (0.6927/0.2954). These results demonstrate that while bagging and sampling already provide substantial gains over the baseline, incorporating the strategy gradient in ELAG further enhances stability and predictive performance, particularly in scenarios of limited sample sizes.

### ELAG enhances prediction and provides biological insights for complex human diseases

To validate ELAG in real-world settings, we applied it to three complex neuro-immune disorders from the UK Biobank: Alzheimer’s disease (AD), rheumatoid arthritis (RA), and multiple sclerosis (MS). Following standard QC, the datasets contained 1,673,294 variants and 4,227 individuals for AD, 1,775,223 variants and 2,008 individuals for RA, and 1,712,010 variants and 3,258 individuals for MS. All of the datasets had an equal number of cases and controls. The Manhattan plots corresponding to the three diseases are shown as Supplementary Figure 1. We benchmarked a GWAS-informed^42-44^ ELAG model (Supplementary Table 7 and Supplementary Note 1.2.1) against a *P*-value thresholding baseline and two wrapper methods (ACO^45^, GA^46^), with all performance metrics reported on a held-out test set.

### ELAG consistently outperforms alternative feature selection methods

Across all three diseases, ELAG demonstrated substantially and consistently superior predictive performance (Figure 3A and 3B and Table 1). For RA, ELAG achieved an AUROC of 0.7354, an improvement over the range of other methods (0.6241–0.6866). For AD, ELAG’s AUROC of 0.7237 surpassed the 0.6699–0.6841 of competitors. Similarly, for MS, ELAG raised the AUROC to 0.7185 from a PTA baseline of 0.6556. These gains were mirrored across other key metrics. For instance, in AD, ELAG’s MCC of 0.351 represented a gain of up to 0.0968 over alternatives, while its F1-score reached 0.7034, driven by enhanced sensitivity in case detection.

**Figure 3.**
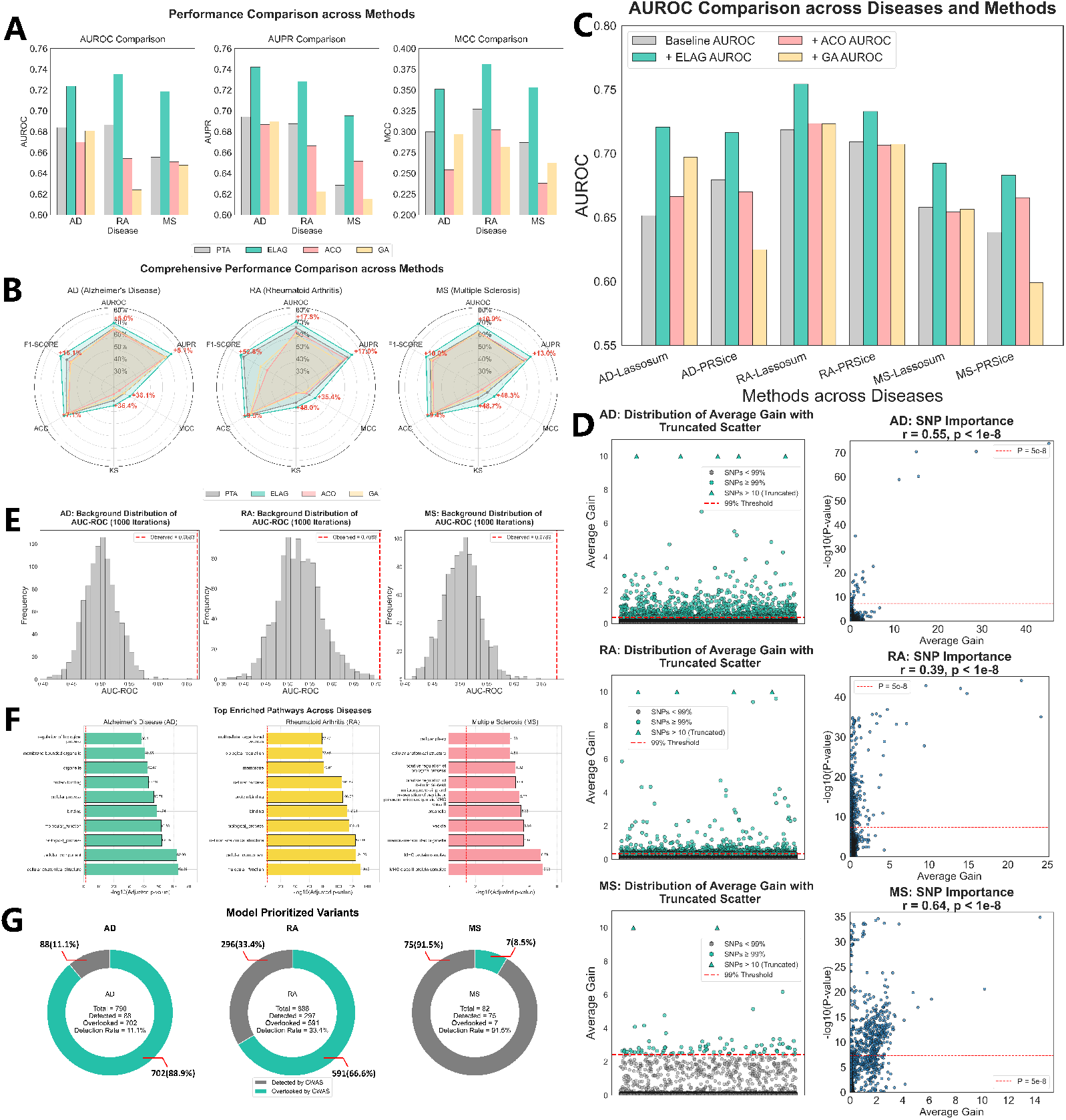
ELAG performance and biological interpretation on three complex diseases. **A**,**B**. ELAG consistently outperforms baseline methods (PTA, ACO, GA) in predicting Alzheimer’s disease (AD), rheumatoid arthritis (RA), and multiple sclerosis (MS), as shown by bar plots of AUROC, AUPR, and MCC (**A**) and summarized in radar plots (**B**). Red numbers denote the relative performance gain of ELAG compared to the lowest-performing method. **C**. Variant sets prioritized by ELAG boost the predictive performance of two standard PRS tools, Lassosum and PRSice, across all three diseases. **D**. Distribution of average gain values for the top-ranked variants in each disease and their relationship with –log_10_(*P*) values from GWAS, highlighting complementary signals. **E**. The predictive power (AUROC) of the top 1% of ELAG-selected variants (red line) is highly significant compared to the null distribution from 1,000 randomly selected variant sets of the same size. **F**. Top biological pathways enriched among genes associated with the highest-ranking ELAG variants, revealing disease-relevant mechanisms for AD, RA, and MS. **G**. A substantial fraction of top 1% ELAG-prioritized variants showed nonsignificant GWAS *P*-values, highlighting ELAG’s ability to capture signals that may be missed by conventional association analyses.

To simulate clinically relevant scenarios with imperfect data, we evaluated ELAG’s robustness to increasing levels of individual genotype missingness (from 20% to 80%) in the test set. The framework showed remarkable resilience. Even at 20% missingness, the drop in AUROC was minimal (AD: -0.0068; RA: -0.0008; MS: -0.0015). Critically, even when faced with 80% missing data, ELAG still retained over 95% of its original AUROC across all three diseases (AD: 0.6983; RA: 0.7069; MS: 0.6879). This stability underscores ELAG’s potential for deployment in real-world clinical applications where genomic data may be incomplete (Supplementary Table 8 and Supplementary Note 1.2.2).

### ELAG variant sets boost the performance of standard polygenic risk scores

We next evaluated ELAG’s capacity to function as a modular feature selection engine for two PRS methods, PRSice and Lassosum. By using the variant subsets identified by ELAG as input for these tools, we observed significant performance gains compared to their standard application across all tested diseases (Figure 3C and Table 2). Specifically, the integration of ELAG with Lassosum for AD increased the AUROC from a baseline of 0.6514 to 0.7207, approaching the performance of the ELAG-only model (0.7237). Similar improvements were observed for MS), where the ELAG-Lassosum model improved the AUROC from 0.6579 to 0.6925 (ELAG-only: 0.7185). Notably, for RA, the synergistic effect was even more pronounced: the ELAG-Lassosum model achieved an AUROC of 0.7543, which not only surpassed the Lassosum baseline (0.7186) but also exceeded the performance of the ELAG-only model (0.7354). In contrast, variant sets generated by alternative algorithms (ACO and GA) provided negligible or even detrimental effects on PRS performance. These results suggest that ELAG’s adaptive, performance-driven selection strategy can identify a more potent and generalizable set of variants, thereby enhancing the predictive power of established genomic tools.

### ELAG identifies compact, predictive, and potentially biologically relevant variant sets

To interpret the models, we analyzed the variants prioritized by the XGBoost ensemble for each disease, ranking them by their average feature importance (‘gain’)^47,48^. We found that a remarkably compact subset of these variants could recapitulate the full model’s performance. For AD, the top 790 variants (top 1%) achieved an AUROC of 0.6683, recovering 92.3% of the full model’s performance. For RA and MS, the top 1% of variants recovered over 96% and 95% of the full model’s AUROC, respectively (Table 3). This high performance was statistically significant, vastly exceeding that of 1,000 randomly selected variant sets of equivalent size (*P* < 10^−3^ for all diseases, Figure 3E).

Crucially, these predictive variant sets were enriched for disease-relevant biology (Figure 3F) using g:Profiler^49^. For AD, prioritized genes converged on pathways including ‘MHC protein complex binding’ and the ‘adaptive immune system,’ both implicated in AD-related neuroinflammation^50,51^(more details can be seen in Supplementary Table 9). For RA, we observed strong enrichment for ‘antigen processing and presentation via MHC class II’ and ‘cytokine-mediated signaling’—vital pillars of RA pathogenesis^52,53^(more details can be seen in Supplementary Table 10). Similarly, for MS, top variants were linked to ‘MHC class II receptor activity’ and ‘T-cell activation,’ aligning with its autoimmune etiology^54,55^(more details can be seen in Supplementary Table 11). For full description can be seen in Supplementary Note 1.2.3.

A key finding was ELAG’s ability to prioritize variants with modest GWAS *P*-values that would be missed by standard filtering (Figure 3G). Within the top 1% of prioritized variants, we identified 702 variants in AD, 591 in RA, and 7 in MS, spanning multiple genes and intergenic regions—all of which may not reach genome-wide significance in GWAS. Representative examples include: *rs9272320* (*HLA-DQA1*; ELAG rank=10 vs. GWAS *P*=0.0189, rank=35,512), implicated in AD neuroimmunity^56^; *rs3130571* (*PSORS1C1*; ELAG rank=27 vs. *P*=0.00340, rank=9,539) in RA^57^; and *rs9274440* (*HLA-DQB1*; ELAG rank=24 vs. *P*=0.00220, rank=6,449) in MS^58,59^. These results may suggest that ELAG may effectively uncover potential variants whose contributions arise from non-additive or complex effects not captured by marginal statistics, though the true functional roles of these variants require further validation. By providing a principled way to discover such hidden signals, ELAG not only improves prediction but may also offer a powerful engine for generating novel, testable biological hypotheses. Full variant lists are provided in Supplementary Tables 12-14.

### Incorporation of non-genetic covariates

To further showcase ELAG’s versatility and robust capability, we investigated its capacity to integrate diverse non-genetic covariates with adaptively selected genetic variant subsets. Utilizing Alzheimer’s disease (AD) as a compelling case study, where age is a well-established and critical determinant, we observed that genetic variants alone yielded an AUROC of 0.7237. Strikingly, upon the incorporation of age at recruitment (a practical surrogate for the generally unrecorded exact onset age) and sex, ELAG’s predictive performance significantly improved, achieving an AUROC of 0.8658 (Supplementary Figure 2, Supplementary Table 15, and Supplementary Note 1.2.4). These findings underscore ELAG’s exceptional ability to effectively synthesize heterogeneous risk factors, leading to enhanced prediction accuracy and broadening its potential applications in comprehensive disease risk modeling.

## Discussion

In this work, we introduced ELAG, a machine learning framework that reframes genomic feature selection from a static filtering problem into a dynamic, adaptive learning task. The central innovation of ELAG is its PG-driven sampling strategy, which learns to select variants by directly optimizing for predictive performance. This approach breaks the reliance on coarse, static *P*-value thresholds that constrain conventional methods. By creating a closed feedback loop where an ensemble of classifiers guides the sampling policy, ELAG can effectively explore the genomic space to uncover a richer, more potent set of disease-associated variants, including those with subtle, non-additive, or context-dependent effects that are systematically missed by standard pipelines.

The power of this adaptive framework was borne out in our extensive evaluations. In simulations designed to mimic the challenging aspects of genomic data—low heritability^60^, small sample sizes^61^, and high polygenicity^62^—ELAG consistently and substantially outperformed the PTA baseline, with average gains of ΔAUROC = 0.0644 and ΔMCC = 0.106. Analysis of feature diversity further revealed low Jaccard similarity (*J* < 0.1) among ensemble members, indicating that ELAG explores complementary feature subspaces and thereby captures signals beyond those prioritized by marginal *P*-value filtering. This simulated advantage translated directly to real-world performance on three complex neuro-immune disorders (AD, RA, and MS). ELAG substantially improved standalone prediction performance (e.g., AUROC for RA increased from 0.6866 to 0.7354) and further enhanced established PRS methods (e.g., Lassosum AUROC for RA from 0.7186 to 0.7543). These consistent gains validate ELAG’s core premise: that a performance-guided, adaptive search may enrich biologically relevant, sub-threshold signals beyond the reach of conventional thresholding.

Beyond its predictive performance, ELAG is designed for real-world clinical utility. A key requirement for real-world deployment is the ability to generate reliable predictions under imperfect data conditions and heterogeneous sources of information. In our analyses, ELAG demonstrated strong robustness to high levels of genotype missingness, a common challenge in clinical datasets, indicating its capacity to deliver stable risk estimates even when genomic data are incomplete. Moreover, its extensible framework enables the seamless incorporation of clinically relevant covariates (e.g., age and sex) alongside adaptively selected genetic variants, thereby enhancing predictive performance potentially and capturing disease risk more comprehensively. Taken together, these properties suggest that ELAG is well-suited for translational applications where both data quality and multimodal integration are critical for precision risk prediction.

Furthermore, ELAG serves as not just a predictive tool but also a potential engine for biological discovery. The compact, high-performance variant sets it identifies are enriched for disease-relevant biological pathways, suggesting that the model may capture biologically meaningful signals. Critically, some of these predictively important variants lack genome-wide significance in standard GWAS, underscoring ELAG’s ability to computationally recover candidate causal variants that may be obscured by stringent multiple-testing corrections in association studies. By prioritizing variants based on their contribution to predictive performance rather than solely on marginal statistical effects, ELAG may provide a principled avenue for generating testable hypotheses about the complex genetic architecture of disease.

Our study has several limitations that suggest future directions. First, while ELAG highlights statistically and biologically plausible variants, their causal roles require experimental validation. Second, its ensemble design, though key to robustness, is more computationally demanding than single-model methods; algorithmic and parallelization advances could improve scalability. Third, this work is an initial attempt to apply PG ideas to genomic feature selection, with scope for refining the policy-learning strategy to enhance efficiency and interpretability. Finally, although we incorporated GWAS and functional annotation priors, the framework is readily extensible to multi-modal data, which may further improve predictive performance and biological insight.

In conclusion, ELAG establishes a powerful new paradigm for polygenic risk prediction by synergistically integrating ensemble learning with PG optimization. By moving beyond static filtering, it mitigates a core bottleneck in genomic analysis. Its demonstrated success in both challenging simulations and real-world disease cohorts paves the way for a new class of intelligent, adaptive models at the intersection of machine learning and precision medicine.

## Methods

### The ELAG framework

#### Overall architecture

ELAG is a machine learning framework designed for polygenic risk prediction from high-dimensional genomic data. The workflow begins with genotype data that has undergone standard quality control to preserve a large variant pool. ELAG’s core is a PG-inspired algorithm that dynamically samples variant subsets, which are then used to train an ensemble of base classifiers. The collective performance of this ensemble provides a reward signal that iteratively refines the sampling policy. The final output is a trained ensemble model for predicting binary disease status (case/control).

The training process is embedded within a five-fold cross-validation scheme to ensure robust evaluation. In each fold, the data is partitioned into an 80% training set and a 20% validation set. Within each fold, the framework executes multiple rounds of policy-guided sampling and model training. Specifically, for each round, a subset of variants is sampled based on the current policy, and a new base classifier is trained on this subset. The predictions from all trained classifiers in the ensemble are averaged, and the resulting binary cross-entropy loss on the validation set is used to compute the policy gradient. This gradient informs the update of the policy parameters, creating a closed loop where predictive performance directly guides feature selection. The final model selected for testing is the ensemble that achieves the highest AUROC on the validation data across all training rounds.

#### Novel policy gradient-inspired algorithm

As illustrated in Figure 1, ELAG assigns each variant a sampling probability *pro*_*i*_based on Equation (1), which reflects its combined importance derived from external information.

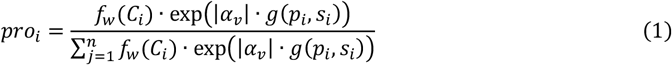

where |α_*v*_| acts as an adaptive temperature value with *v* corresponding to the cross-validation fold, which modulates the sharpness of the sampling distribution. It is dynamically updated according to classification performance, thereby adjusting the selection bias over variants. *p*_*i*_ represents the *P*-value of variant *i* and *s*_*i*_ denotes the corresponding *P*-value for variant *i* from external GWAS summary statistics information. The function *g*(*p*_*i*_, *s*_*i*_) represents a trainable combination of *p*_*i*_ and *s*_*i*_, parameterized by a learnable weight *β*:

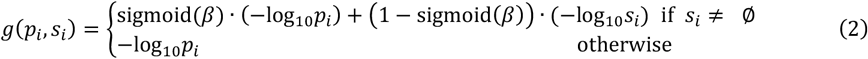

where *β* serves as a tunable scalar that balances the contribution of external GWAS-derived signals (e.g., *P*-values) and annotation scores (e.g., CADD). The sigmoid function constrains *β* to the interval (0, 1), ensuring a balance between data sources. When annotation scores are available, *C*_*i*_ represents the corresponding annotation score for that variant *i. f*_*w*_(*C*_*i*_) is a weighting function based on the input *C*_*i*_, the annotation score for that variant *i*; its explicit form is given by

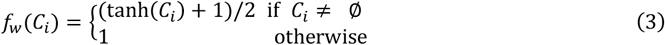

For backpropagation,we establish the following gradient equation (4) to update the parameters in the framework through loss.

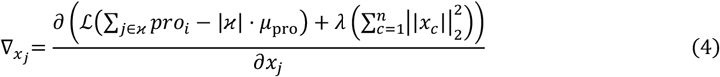

where the gradient is computed as 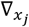, where *x*_*j*_ represent a learnable parameter. ϰ denotes the set of variants selected in the current iteration, ℒ is the loss value, *λ* is a tunable hyperparameter, and *n* is the number of learnable parameters. *μ*_*pro*_ denotes the average probabilities assigned to all variants.

The gradient 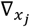 is constructed such that subsets of variants whose combined sampling probability ^∑^_*j*∈ϰ_ *pro*_*i*_exceeds the global average |ϰ| · *μ*_*pro*_ are preferentially reinforced in proportion to their impact on the binary cross-entropy loss ℒ. This design helps to mitigate fluctuations caused by varying batch sizes and ensures stability during optimization. L_2_ regularization term 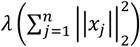 imposes weight decay to improve generalization. Furthermore, by operating on the raw sum of probabilities ∑_*j*∈ϰ_ *pro*_*i*_ rather than the log-sum ∑_*j*∈ϰ_ l*o*g(*pro*_*i*_), this formulation avoids numerical instability caused by extremely small probability values. When the probability vector is derived from Equation (1) over more than 1,000,000 variants, the resulting probabilities can become exceedingly small. Applying a logarithm to such values would produce large negative numbers, leading to unstable gradients and potential exploding loss values. By using the raw sum, we maintain both numerical stability and a meaningful signal for guiding the adaptive selection of informative variant subsets in high-dimensional settings.

Thus, the parameters can be updated by gradient descent, as follows:

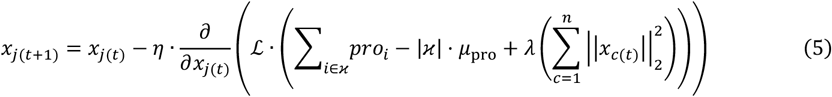

Equation (5) defines the update rule that adjusts the learnable parameters from iteration *t* to *t*+1 based on the computed gradient and the learning rate *η*.

### Incorporation of non-genetic covariates

ELAG is designed primarily for genomic data but readily accommodates non-genetic covariates (e.g., clinical phenotypes) when they are relevant to disease. Inclusion of non-genetic covariates is optional and not the primary focus of this study. For the real-world analyses, we concatenated covariates—specifically age at recruitment and sex—with the variant subsets sampled by ELAG; the combined feature vectors were then used jointly for classifier training and evaluation. For sample *i*, let sex_*i*_ denote sex (encoded as 0 = female, 1 = male), age_*i*_ denote age at recruitment, and *G*_*i*1_, …, *G*_*in*_ denote the *n* variant features collected for that sample. Each ELAG-sampled variant subset was concatenated with the covariates as

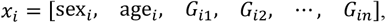

and these combined vectors were used for classifier training and subsequent evaluation.

### Model training and testing

The genomic dataset was randomly divided into a training set (90%) and an independent testing set (10%). A 1:1 case-to-control ratio was maintained in all datasets to ensure balanced class representation during model training and evaluation.

We implemented five machine learning classifiers: AdaBoost, Naive Bayes, CatBoost, GBDT, and XGBoost. Hyperparameter tuning was conducted using grid search based on the scikit-learn (v1.3.0) framework. The models yielding the highest AUROC on the validation sets were selected for final evaluation.

Our proposed policy gradient–inspired optimization algorithm was implemented in PyTorch (v2.0.1). The learning rate was set to 0.1, with an L_2_ regularization coefficient (*λ*) of 0.01. The models were trained for 20 iterations to ensure performance stability.

For both ACO^45^ and GA^46^, we wrapped XGBoost–based AUROC on a validation split as the fitness function and used grid search to identify optimal hyperparameters (population size, generations, crossover/mutation rates for GA; number of ants, iterations, pheromone evaporation, α/β weights for ACO). Due to the exponential growth of the combinatorial search space and resulting computational constraints, we first filtered variants by GWAS *P* < 0.01, reducing the candidate pool to 19,231 variants for AD, 21,101 for RA, and 21,320 for MS before applying these wrapper methods.

### Polygenic Risk Score Methods

In this study, we applied two widely used PRS approaches—PRSice^40^ and Lassosum^41^—as our genetic risk predictors. To integrate these with ELAG, we used ELAG as a feature selector. In our project, ELAG was trained using cross-validation, and in each fold, multiple bagged classifiers were generated, each selecting its own subset of variants. For example, with 5-fold cross-validation and 10 classifiers per fold, this resulted in 50 distinct feature sets across all folds.

We then passed each of these ELAG-selected variant sets into the PRS pipelines. For each PRS method, scores were computed on the test set separately for each feature set. The resulting 50 PRS scores were then averaged to produce a single ELAG–enhanced PRS prediction for each individual (Supplementary Figure 3).

As a baseline, we also computed PRS using the full genome-wide variant set, without ELAG-based feature selection, to assess the added predictive value of integrating ELAG with standard PRS methods.

### Model interpretation

In order to evaluate the contribution of each SNP feature to the predictive performance of the XGBoost models, we adopted the gain metric^47,48^, which quantifies the average improvement of the objective function when a feature is used for a split. Specifically, for a given split, the gain is defined as:

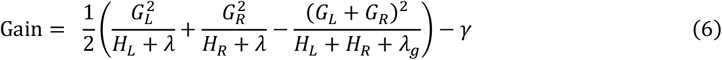

where *G*_*L*_ and *G*_*R*_ denote the sums of the first-order gradients in the left and right child nodes, respectively, while *H*_*L*_ and *H*_*R*_ denote the corresponding sums of the second-order gradients. The term *λ*_*g*_ and is the L2 regularization coefficient, and *γ* is the minimum split loss parameter that penalizes overly complex trees. Intuitively, the gain measures the reduction in the loss function after a split compared with the parent node, adjusted by regularization. The overall feature importance of each SNP was obtained by aggregating the gains across all occurrences of that feature in all trees and reporting their average values, following the default definition of importance_type=‘gain’ in XGBoost.

After training an ensemble of classifiers, we ranked all variants by their gain and discarded those with zero gain, retaining only the top 1% of variants. We then assessed the predictive power of this subset by retraining the classifiers on the training data using only these top variants and evaluating performance on the held-out test set. As a null benchmark, we performed the same procedure 1,000 times with randomly sampled variants of equal cardinality, training and testing under identical conditions. Empirical *P*-values were estimated via Monte Carlo^63^, defined as

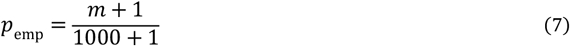

where *m* is the number of random replicates whose background AUROC met or exceeded the observed AUROC. For the three diseases studied, the total number of variants involved was 790 for AD, 888 for RA, and 82 for MS. To investigate the biological functions associated with the genes linked to these variants, we performed functional enrichment analysis using g:Profiler^49^. Analyses included Gene Ontology (GO)^64^, Human Protein Atlas (HPA)^65^, and Reactome (REAC)^66^ pathways.

Furthermore, we also quantified how many of our top-ranking variants may be missed by a standard GWAS threshold. Specifically, we first took the top 1% of variants by XGBoost gain importance and, for each of those variants, retrieved its GWAS *P*-value. We then sorted these *P*-values in ascending order to assign each variant a rank. Next, we compared the rank of each variant against the optimal feature set size identified by our grid search (the number of variants that maximized classifier performance). Any variant whose *P*-value rank exceeded that optimal set size—meaning it would not have been selected by a hard cutoff at that rank— was deemed potentially overlooked by conventional association analysis.

### Jaccard similarity

To quantify the overlap between different sets of selected QTLs, we employed the Jaccard similarity coefficient. For any two sets *A* and *B*, the Jaccard index is defined as

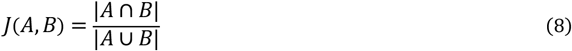

where |*A* ∩ *B*| is the number of elements common to both sets and |*A* ∪ *B*| is the total number of unique elements across both sets. A higher Jaccard index indicates greater agreement between two selection methods or replicates, whereas a lower value reflects more divergent QTL choices.

### Simulation data

Genotypes for 2,596 variants on chromosome 1 were simulated based on allele frequencies and linkage disequilibrium (LD) coefficients from the European (EUR) panel of the 1000 Genomes Project^67^. Genotypes were encoded as 0, 1, or 2, representing the number of alternative alleles. Only variants with a minor allele frequency (MAF) above 5% and pairwise LD *r*^2^<0.1 were retained to ensure low redundancy.

Given the genotypes, subjects’ and phenotypes were simulated using a liability-threshold model^68,69^. The relationship between multiple variant genotypes and liability are present in the following equation (9), a linear–nonlinear mixed model (LNMM).

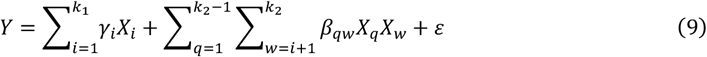

where the effect sizes *γ* and *β* were drawn from a uniform distribution between 0 and 1, with X denoting the genotype of a given variant. We set *k*_1_ and *k*_2_, corresponding to the numbers of linear and nonlinear QTLs, respectively, where *k*_1_=*k*_2_=500. The environmental component ε followed a normal distribution *N*(0, σ^2^).

For each individual, the total genetic liability was computed as the sum of the effects of alternative alleles at causal QTLs, combined with a random environmental contribution. The variance of environmental noise was calibrated to achieve a targeted heritability *h*^2^ (0.2, 0.3, 0.4). A synthetic population of 3 million individuals was generated. Individuals in the top 1% of the liability distribution were labeled as cases, simulating a disease prevalence of 1%; the rest were treated as controls. From this population, case/control cohorts of sizes 500, 1,500, and 5,000 were randomly sampled without replacement for downstream analysis.

Functional annotation scores in simulations were generated under two conditions: (1) informative annotations, where true QTLs received scores drawn from *N* ( *μ* = 0.75, σ^2^ = 0.1) and non-QTLs from *N*(*μ* = 0.25, σ^2^ = 0.1); and (2) shuffled annotations, in which those same scores were randomly permuted across variants. All simulated scores were truncated to the [0, 1] interval.

### Real data: UK Biobank

All genomic data and phenotypes in the real datasets are from the UK Biobank with the ICD-10 diagnoses. All individuals included in the study were born in the UK. The AD, RA, and MS sub-datasets have sample sizes of 4,227, 2,008, and 3,258, respectively. The case-to-control ratio is maintained at 1:1 in each of the three datasets.

Variants were preprocessed and analyzed by PLINK 2.0 ^70^ and KGGA (https://pmglab.top/kgga/)^71^ based on the following criteria: (1) subject call rate ≥ 98%; (2) variant call rate ≥ 98%; (3) Hardy-Weinberg equilibrium *P*-value ≥ 1 × 10^−5^; (4) MAF ≥ 5%; (5) LD pruning threshold *pp*^2^ < 0.9. The model focused on autosomal chromosomes. After applying standard quality control, the number of variants per sample is 1,673,294 for AD, 1,775,223 for RA, and 1,712,010 for MS.

### Computational implementation

The computational experiments were carried out on a dedicated desktop workstation configured as follows: 32 GB of dual-channel GLOWAY DDR5 RAM clocked at 6000 MHz; an Intel^®^ Core™ i7-13700K CPU (3.4 GHz base, up to 5.4 GHz turbo); a 2 TB PCIe 4.0 NVMe SSD (aigo P7000Z series); and an NVIDIA GeForce RTX 4090 GPU with 24 GB of GDDR6X memory. As an illustrative example in the Alzheimer’s disease case study, a single CPU core dynamically sampled 5,000 variants per classifier (5-fold cross-validation repeated 10 times, totaling 50 independent models) from a pool of 1,673,294 variants. Under these conditions, each training epoch required approximately 5.9 minutes of wall-clock time and incurred an average additional memory overhead of 593.97 MB.

## Data availability

In this study, partial genotype data from the UK Biobank (https://www.ukbiobank.ac.uk/) was accessed through a collaboration with application no.86920. Whole Exome Sequencing (WES) data in VCF.GZ format was accessed under Field ID 23157 and imputed genotypes in PGEN format were accessed under Field ID 22828. Data are available for bona fide researchers upon application to the UK Biobank, and the dataset of the 1000 Genomes Project is shown at https://www.internationalgenome.org/, which does not require access rights.

## Code availability

The source code of ELAG is publicly available at https://github.com/LHDLHUB/ELAG.

## Data Availability

All data produced in the present study are available upon reasonable request to the authors.

https://github.com/LHDLHUB/ELAG

## Supplementary information

### Supplementary Information

Supplementary Note, Figs. 1-3, and Tables 1-8, 15.

### Supplementary Tables

Supplementary Tables 9-14.

