## Supplementary Information. for "Policy gradient-guided ensemble learning for enhanced polygenic risk prediction in ultra-high-dimensional genomics"

#### Catalogue

### 1 Supplementary notes

#### 1.1 Simulations: ELAG compatibility with multiple machine learning classifiers

To assess ELAG's compatibility with diverse predictive algorithms, we benchmarked its performance against baseline feature selection across five representative machine learning classifiers—XGBoost, AdaBoost, Naïve Bayes, CatBoost, and Gradient Boosting Decision Trees (GBDT)—under identical simulation settings (number of QTLs = 1,000, heritability $h^2$=0.2, sample size = 3,000), as shown in Supplementary Table 3.

Across all classifiers, ELAG consistently outperformed the baseline in key discrimination metrics. For example, with XGBoost, ELAG achieved an AUROC of 0.7082 versus 0.6585 for the baseline, and an AUPR of 0.6893 versus 0.6162, representing relative gains of 7.5% and 11.9%, respectively. Similar improvements were observed in MCC (0.3556 vs. 0.2802) and KS statistic (0.3667 vs. 0.2800). In AdaBoost, the AUROC improvement was more modest (0.6468 vs. 0.6380), yet ELAG still showed higher KS (0.2667 vs. 0.2467) and MCC (0.2471 vs. 0.2400), indicating consistently better separation between case and control distributions.

The most pronounced gains were seen in GBDT, where ELAG improved AUROC from 0.6442 to 0.7125 (+10.6%) and AUPR from 0.6052 to 0.7003 (+15.7%), with corresponding MCC improvement from 0.2610 to 0.3467. Naïve Bayes and CatBoost also exhibited robust benefits: for Naïve Bayes, ELAG raised AUROC from 0.6828 to 0.7135 (+4.5%) and AUPR from 0.6807 to 0.7226 (+6.1%), while for CatBoost, AUROC increased from 0.6420 to 0.6910 (+7.6%) and MCC from 0.2317 to 0.3472 (+49.8%).

These results highlight three important trends. First, ELAG's improvements may be consistent across heterogeneous learning paradigms. Second, the magnitude of gain is classifier-dependent, with greater relative boosts in models (e.g., CatBoost, GBDT). Third, even in classifiers where baseline performance is already strong, ELAG's adaptive sampling guided by policy gradient confer additional benefits in calibration and robustness. Collectively, these findings may underscore ELAG's model-agnostic applicability and its potential as a general-purpose enhancement for genomic risk prediction pipelines.

#### 1.2 Real-word data application

##### 1.2.1 Using ELAG

Owing to its extensible architecture, ELAG can seamlessly integrate external sources of variant-level information—such as GWAS summary statistics and functional annotations (e.g., CADD scores)—into its prioritization framework. This flexibility allows users to tailor the feature set to the available resources and disease context. We evaluated ELAG's performance in three real-world disease cohorts—Alzheimer's disease (AD), rheumatoid arthritis (RA), and multiple sclerosis (MS)—under two configurations: using GWAS statistics alone (ELAG+GWAS) and using both GWAS statistics and CADD scores (ELAG+GWAS+CADD) (Supplementary Table 7).

Across all diseases, ELAG+GWAS generally yielded slightly higher or comparable performance relative to ELAG+GWAS+CADD, suggesting that while functional scores can be incorporated, their predictive contribution is context-dependent. For AD, AUROC was 0.7237 for ELAG+GWAS and 0.7199 for ELAG+GWAS+CADD, with AUPR showing a negligible difference (0.7421 vs. 0.7450). RA showed a similar trend (AUROC: 0.7354 vs. 0.7214), while MS presented the smallest gap (AUROC: 0.7185 vs. 0.7104). In all cases, MCC values remained stable, and no significant degradation in classification metrics was observed upon adding CADD scores.

This pattern aligns with our simulation findings: ELAG preserves predictive stability despite the inclusion of auxiliary features with uncertain or heterogeneous utility. Specifically, the incorporation of CADD—despite its known biases toward certain functional categories—did not cause systematic performance deterioration. Instead, the model maintained stability in discrimination (AUROC, AUPR) and calibration-related metrics (MCC, KS), underscoring its resilience to heterogeneity in feature informativeness.

Collectively, these results indicate that ELAG may have potential to accommodate diverse genomic annotation layers without compromising baseline predictive performance, thereby enabling flexible adaptation to varying levels of annotation richness in clinical and research settings.

##### 1.2.2 The robustness of ELAG against missingness

To assess ELAG's robustness under realistic clinical scenarios with incomplete genotype data, we systematically evaluated its performance across per-individual genotype-level missingness rates of 20%, 40%, 60%, and 80% in three diseases. For each missingness level, we randomly masked the corresponding proportion of genotypes in the test set and directly applied the pre-trained ELAG model without imputation.

In the absence of missingness, ELAG achieved strong baseline performance (AD: AUROC = 0.7237, RA: 0.7354, MS: 0.7185), outperforming comparator methods such as PTA, ACO, and GA across all metrics. With increasing missingness, performance degradation was gradual rather than abrupt. For example, in AD, AUROC decreased only modestly from 0.7237 (0%) to 0.7068 (40%) and 0.6983 (80%), with AUPR remaining consistently above 0.72. Similarly, RA maintained AUROC values above

0.7069 even at 80% missingness, and MS retained AUROC = 0.6879 at the highest dropout level. Notably, in several cases (e.g., RA at 20% and 40% missingness), recall remained high (>0.89 at 40% missingness), reflecting preserved sensitivity despite data loss.

These results indicate that ELAG's architecture—leveraging feature selection within the ELAG framework and the inherent robustness of tree-based learners—confers resilience to extensive locus dropout. Even when confronted with extreme missingness (80%), ELAG retained predictive metrics above comparator baselines in the complete-data setting, underscoring its potential for deployment in real-world clinical pipelines where genotype completeness cannot be guaranteed.

##### 1.2.3 Enrichment analysis of genes in the three diseases

To explore the biological meaning of the variants prioritized by ELAG, we performed pathway and tissue-enrichment analysis on genes implicated by the top 1% of aggregated XGBoost importance gains for each disease using g:Profiler. We focused on Gene Ontology[1] categories (BP, MF, CC), Reactome[2] (REAC) pathways and Human Protein Atlas[3] (HPA) tissue annotations; full enrichment tables are provided in Supplementary Tables 9–11.

Across all three disorders the enrichment profile consistently implicated immune and nervous-system related processes, providing convergent support for ELAG's ability to recover biologically plausible signals from large feature sets:

AD: Top enrichments included MHC-related molecular functions[4] (GO:0023023, "MHC protein complex binding") and adaptive immunity pathways[5] (Reactome: R-HSA-1280218, "Adaptive Immune System"), together with signal transduction[6] (GO:0007165) and hippocampus-specific[7] expression (HPA:0250000). These annotations are consistent with growing evidence that immune processes and hippocampal biology contribute to AD pathogenesis, and indicate that ELAG-prioritized loci are enriched for genes with putative roles at the neuro-immune interface.

RA: Enriched categories included antigen processing and presentation[8] (GO:0019882), cytokine-mediated signaling[9] (GO:0019221), and neuronal signaling/development terms[10] (GO:0023041, GO:0007399), as well as cerebellum tissue[11] annotations (HPA:0090000). The immune pathway enrichments are related to RA's established autoimmune etiology, while the neuronal terms point to potential neuro-immune or neuroinflammatory components captured by ELAG in the prioritized gene set.

MS: Key enrichments included MHC class II receptor activity[12] (GO:0032395), T-cell activation[13] (GO:0042110), and neuron development[14] (GO:0048666), together with broader cellular regulation[15] (GO:0048522) and tissue annotations for the cerebral cortex[16, 17] (HPA:0100000) and lung[18, 19] (HPA:0300000). The prominence of antigen-presentation and T-cell activation pathways is concordant with established MS pathophysiology and supports the biological relevance of ELAG's prioritized loci. Tissue enrichments for the cerebral cortex are directly consistent with MS as a central-nervous-system disorder; the lung annotation may reflect systemic immune interactions or environmental and comorbidity-related factors and therefore warrants further investigation.

Together, these results suggest that ELAG preferentially highlights loci mapping to genes involved in immune function and neuronal processes—consistent with the concept of a neuro-immune axis[20-23] for these disorders. However, several limitations must be noted. Variant→gene assignment can be ambiguous (e.g., intergenic variants, long-range regulation), and pathway enrichment is influenced by gene-set size and annotation bias. Additionally, enrichment analyses are hypothesis-generating and do not establish causality. We have provided complete enrichment statistics in Supplementary Tables 9–11.

#### 1.2.4 Incororation of non-genetic covariates using ELAG

Supplementary Table 15 provides a comparison of model performance when integrating non-genetic covariates with genetic risk prediction using ELAG. The results robustly demonstrate the broad applicability and utility of our approach across diverse diseases.

The effectiveness of incorporating covariates is context-dependent and aligns with the known etiologies of each disease. For AD, a condition with strong age-dependent risk, the addition of age at recruitment and sex covariates ("All features") led to universal improvement across all metrics compared to genetics alone. This is particularly evident in the increase of AUROC (from 0.724 to 0.866) and MCC (from 0.351 to 0.592), confirming that ELAG successfully captures the critical non-genetic risk factors most relevant to AD. The significant performance gain underscores the value of including age at recruitment for diseases where age is a major proximal risk factor.

For RA and MS, where the influence of age at recruitment and sex is comparatively less pronounced than in AD, the genetic component alone ("Only genetics") already provides a strong predictive signal. Notably, for RA, the genetics-only model even achieved a marginally higher AUROC than the full model (0.735 vs. 0.742). However, a holistic view across the spectrum of performance metrics reveals that the incorporation of these covariates via ELAG still yields a more balanced and robust model. Specifically, the "All features" model consistently maintains high performance across AUPR, Precision, and F1 Score, mitigating potential biases that might arise from relying on a single metric. This underscores a key advantage of ELAG: it allows for the integration of readily available auxiliary variables like age at recruitment and sex, which may enhance model stability and clinical interpretability, even when their standalone predictive power is modest.

In conclusion, the ELAG framework offers a flexible strategy for polygenic prediction. It empowers researchers to leverage core demographic variables such as age at recruitment to tailor risk models, whether they are essential drivers of risk or valuable auxiliary variables that contribute to a more comprehensive and stable risk prediction.

### 2 Supplementary figure

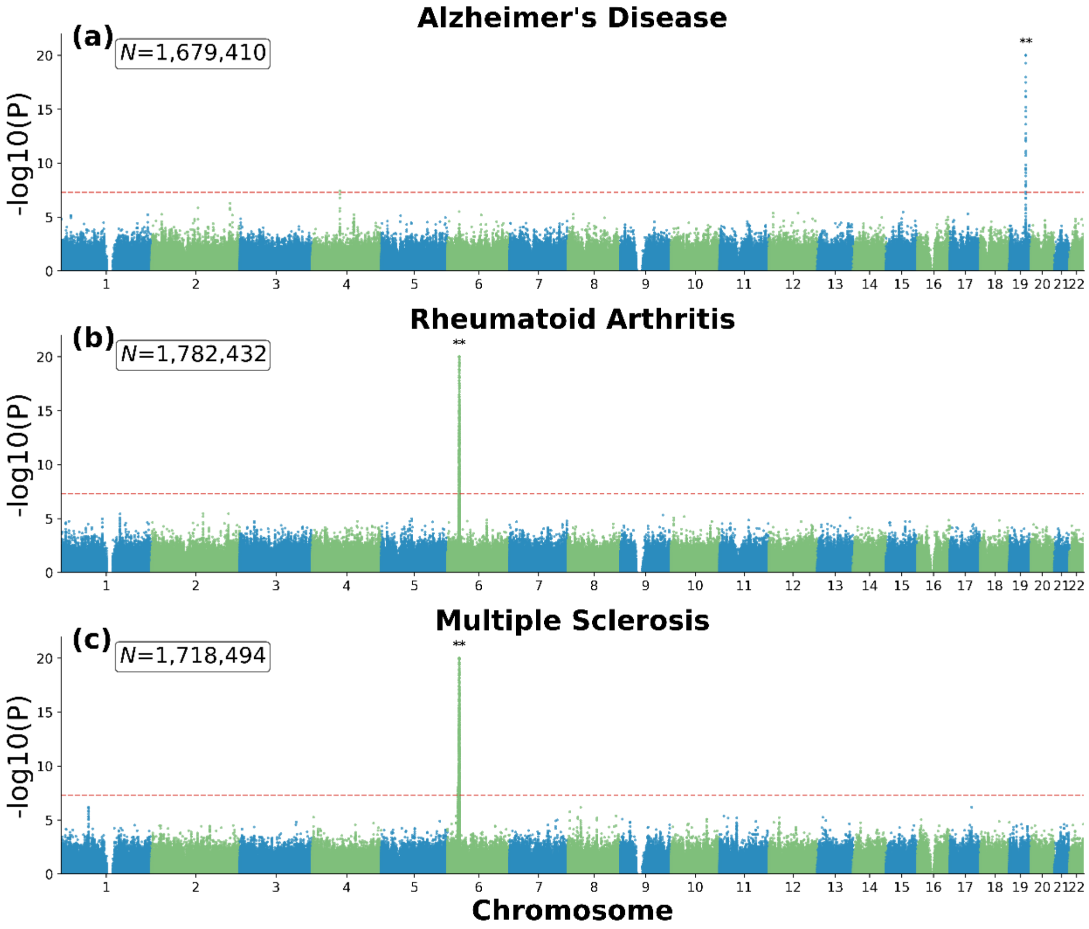

**Supplementary Figure 1. The manhattan plots of three diseases (Alzheimer's disease, rheumatoid arthritis, and multiple sclerosis).**

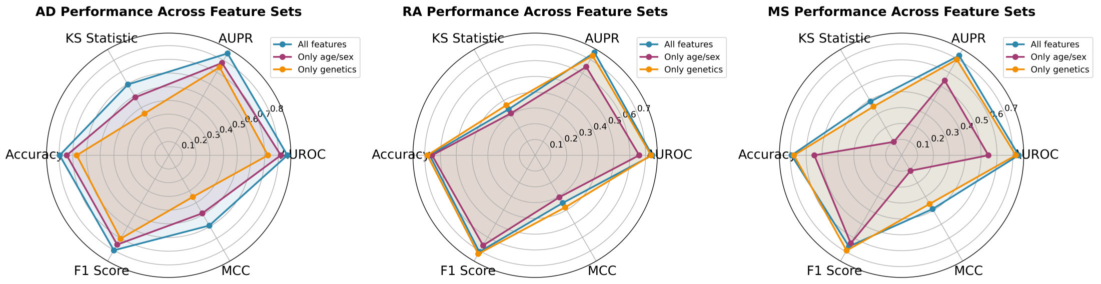

**Supplementary Figure 2. Comparative performance analysis of feature sets across diseases.**

Radar charts display the performance of three feature sets (All features, Only age/sex, Only genetics) across six evaluation metrics for Alzheimer's Disease (AD), Rheumatoid Arthritis (RA), and Multiple Sclerosis (MS).

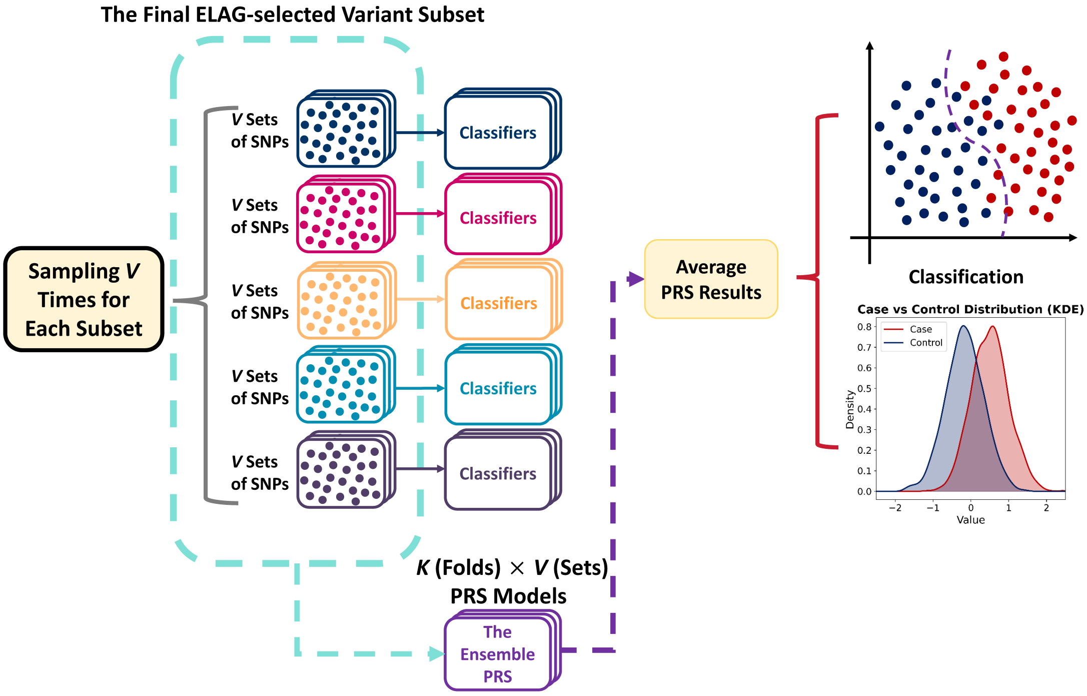

**Supplementary Figure 3. Workflow of integrating ELAG with polygenic risk score methods.**

Genotype data were first processed by ELAG, which, through cross-validation and bagging, generated multiple classifiers, each selecting a subset of variants. After fininshing training, these ELAG-selected variant subsets were then passed into PRS pipelines (Lassosum or PRSice) to compute individual risk scores. The PRS predictions obtained from all subsets were subsequently averaged, yielding a single ELAG–enhanced PRS prediction for each individual.

### 3 Supplementary tables

| Sample Size | $h^2$ | Method | AUROC | AUPR | KS | ACC | Recall | Precision | F1 Score | MCC |
|---|---|---|---|---|---|---|---|---|---|---|
| 1,000 | 0.2 | ELAG | 0.6324 | 0.6405 | 0.2800 | 0.6300 | 0.5000 | 0.6758 | 0.5747 | 0.2693 |
| | | Baseline | 0.5680 | 0.5951 | 0.1600 | 0.5800 | 0.4800 | 0.6000 | 0.5330 | 0.1633 |
| | 0.3 | ELAG | 0.6324 | 0.6243 | 0.2800 | 0.6400 | 0.7400 | 0.6167 | 0.6727 | 0.2858 |
| | | Baseline | 0.5736 | 0.5601 | 0.1800 | 0.5800 | 0.7800 | 0.5571 | 0.6500 | 0.1746 |
| | 0.4 | ELAG | 0.6668 | 0.6834 | 0.3000 | 0.6400 | 0.9400 | 0.5875 | 0.7231 | 0.3500 |
| | | Baseline | 0.5932 | 0.5680 | 0.2400 | 0.6200 | 0.9200 | 0.5750 | 0.7077 | 0.3000 |
| 3,000 | 0.2 | ELAG | 0.6704 | 0.6432 | 0.2933 | 0.6433 | 0.6733 | 0.6352 | 0.6537 | 0.2872 |
| | | Baseline | 0.6308 | 0.6362 | 0.2133 | 0.6067 | 0.6533 | 0.5976 | 0.6242 | 0.2143 |
| | 0.3 | ELAG | 0.7082 | 0.6893 | 0.3667 | 0.6767 | 0.7333 | 0.6587 | 0.6940 | 0.3556 |
| | | Baseline | 0.6585 | 0.6162 | 0.2800 | 0.6367 | 0.7467 | 0.6120 | 0.6727 | 0.2802 |
| | 0.4 | ELAG | 0.7484 | 0.7432 | 0.4067 | 0.6967 | 0.7400 | 0.6810 | 0.7093 | 0.3948 |
| | | Baseline | 0.7275 | 0.7131 | 0.3733 | 0.6833 | 0.6800 | 0.6846 | 0.6823 | 0.3667 |
| 10,000 | 0.2 | ELAG | 0.7180 | 0.6863 | 0.3500 | 0.6730 | 0.7280 | 0.6559 | 0.6900 | 0.3481 |
| | | Baseline | 0.6927 | 0.6629 | 0.2920 | 0.6430 | 0.7680 | 0.6144 | 0.6827 | 0.2954 |
| | 0.3 | ELAG | 0.7576 | 0.7390 | 0.4080 | 0.7000 | 0.7860 | 0.6706 | 0.7238 | 0.4060 |
| | | Baseline | 0.7271 | 0.7122 | 0.3660 | 0.6810 | 0.7540 | 0.6580 | 0.7027 | 0.3659 |
| | 0.4 | ELAG | 0.8206 | 0.8127 | 0.5060 | 0.7490 | 0.7380 | 0.7546 | 0.7460 | 0.4981 |
| | | Baseline | 0.7969 | 0.7902 | 0.4545 | 0.7230 | 0.7780 | 0.7010 | 0.7374 | 0.4487 |

**Supplementary Table 1.** Performance metrics (AUROC, AUPR, KS, ACC, recall, precision, F1, MCC) for ELAG and baseline across sample sizes (1k, 3k, 10k) and heritabilities ($h^2$ = 0.2, 0.3, 0.4). Simulations included 1,000 causal QTLs, with half exhibiting linear effects and half nonlinear effects.

| Sample Size | QTL Number | Method | AUROC | AUPR | KS | ACC | Recall | Precision | F1 Score | MCC |
|---|---|---|---|---|---|---|---|---|---|---|
| 1,000 | 100 | ELAG | 0.6464 | 0.5994 | 0.2600 | 0.6300 | 0.7200 | 0.6101 | 0.6606 | 0.2643 |
| | | Baseline | 0.6356 | 0.6366 | 0.2600 | 0.6300 | 0.5200 | 0.6667 | 0.5843 | 0.2665 |
| | 500 | ELAG | 0.6516 | 0.6730 | 0.2800 | 0.6300 | 0.5400 | 0.6583 | 0.5934 | 0.2643 |
| | | Baseline | 0.6376 | 0.6145 | 0.2800 | 0.6300 | 0.8000 | 0.5970 | 0.6838 | 0.2765 |
| | 1,000 | ELAG | 0.6324 | 0.6405 | 0.2800 | 0.6300 | 0.5000 | 0.6758 | 0.5747 | 0.2693 |
| | | Baseline | 0.5680 | 0.5951 | 0.1600 | 0.5800 | 0.4800 | 0.6000 | 0.5330 | 0.1633 |
| 3,000 | 100 | ELAG | 0.7115 | 0.7038 | 0.3200 | 0.6567 | 0.7733 | 0.6270 | 0.6925 | 0.3222 |
| | | Baseline | 0.7225 | 0.6896 | 0.3667 | 0.6767 | 0.6533 | 0.6853 | 0.6689 | 0.3537 |
| | 500 | ELAG | 0.6949 | 0.6803 | 0.3200 | 0.6600 | 0.7533 | 0.6348 | 0.6890 | 0.3257 |
| | | Baseline | 0.6642 | 0.6471 | 0.2800 | 0.6400 | 0.6800 | 0.6296 | 0.6538 | 0.2810 |
| | 1,000 | ELAG | 0.6704 | 0.6432 | 0.2933 | 0.6433 | 0.6733 | 0.6352 | 0.6537 | 0.2872 |
| | | Baseline | 0.6308 | 0.6362 | 0.2133 | 0.6067 | 0.6533 | 0.5976 | 0.6242 | 0.2143 |
| 10,000 | 100 | ELAG | 0.8102 | 0.7844 | 0.4780 | 0.7390 | 0.8060 | 0.7110 | 0.7554 | 0.4824 |
| | | Baseline | 0.8083 | 0.7834 | 0.4660 | 0.7320 | 0.7460 | 0.7257 | 0.7357 | 0.4642 |
| | 500 | ELAG | 0.7761 | 0.7717 | 0.4200 | 0.7050 | 0.6560 | 0.7273 | 0.6898 | 0.4198 |
| | | Baseline | 0.7789 | 0.7704 | 0.4240 | 0.7120 | 0.7840 | 0.6850 | 0.7313 | 0.4285 |
| | 1,000 | ELAG | 0.7180 | 0.6863 | 0.3500 | 0.6730 | 0.7280 | 0.6559 | 0.6900 | 0.3481 |
| | | Baseline | 0.6927 | 0.6629 | 0.2920 | 0.6430 | 0.7680 | 0.6144 | 0.6827 | 0.2954 |

**Supplementary Table 2.** Performance metrics (AUROC, AUPR, KS, ACC, recall, precision, F1, MCC) for ELAG and baseline methods across varying sample sizes (1k, 3k, 10k) and different QTL numbers (100, 500, 1,000). Simulations included 1,000 causal QTLs, with half exhibiting linear effects and half nonlinear effects.

| Classifiers | Method | AUROC | AUPR | KS | ACC | Recall | Precision | F1 Score | MCC |
|---|---|---|---|---|---|---|---|---|---|
| AdaBoost | ELAG | 0.6468 | 0.6371 | 0.2667 | 0.6233 | 0.5933 | 0.6312 | 0.6117 | 0.2471 |
| | Baseline | 0.6380 | 0.6339 | 0.2467 | 0.6200 | 0.6067 | 0.6233 | 0.6148 | 0.2400 |
| Naïve Bayes | ELAG | 0.7135 | 0.7226 | 0.3667 | 0.6833 | 0.6067 | 0.7165 | 0.6570 | 0.3711 |
| | Baseline | 0.6828 | 0.6807 | 0.2933 | 0.6467 | 0.6670 | 0.6410 | 0.6535 | 0.2936 |
| Catboost | ELAG | 0.6910 | 0.6727 | 0.3533 | 0.6733 | 0.7000 | 0.6646 | 0.6818 | 0.3472 |
| | Baseline | 0.6420 | 0.6261 | 0.2400 | 0.6100 | 0.7667 | 0.5838 | 0.6628 | 0.2317 |
| GBDT | ELAG | 0.7125 | 0.7003 | 0.3600 | 0.6733 | 0.6733 | 0.6733 | 0.6733 | 0.3467 |
| | Baseline | 0.6442 | 0.6052 | 0.2667 | 0.6300 | 0.6733 | 0.6196 | 0.6454 | 0.2610 |
| XGBoost | ELAG | 0.7082 | 0.6893 | 0.3667 | 0.6767 | 0.7333 | 0.6587 | 0.6940 | 0.3556 |
| | Baseline | 0.6585 | 0.6162 | 0.2800 | 0.6367 | 0.7467 | 0.6120 | 0.6727 | 0.2802 |

**Supplementary Table 3.** Performance metrics (AUROC, AUPR, KS, ACC, recall, precision, F1, MCC) for ELAG and baseline methods across different machine learning methods (XGBoost, AdaBoost, Naive Bayes, CatBoost and Gradient Boosting Decision Trees(GBDT)), with a heritability of 0.2 and a sample size of 3,000. Simulations included 1,000 causal QTLs, with half exhibiting linear effects and half nonlinear effects.

| Methods | AUROC | AUPR | KS | ACC | Recall | Precision | F1 Score | MCC |
|---|---|---|---|---|---|---|---|---|
| ELAG | 0.6324 | 0.6405 | 0.2800 | 0.6300 | 0.5000 | 0.6758 | 0.5747 | 0.2693 |
| ELAG+GWAS | 0.7264 | 0.7046 | 0.4000 | 0.6900 | 0.6000 | 0.7317 | 0.6593 | 0.3831 |
| ELAG+GWAS+CADD | 0.7412 | 0.7050 | 0.4400 | 0.7200 | 0.5600 | 0.8230 | 0.6667 | 0.4644 |
| ELAG+GWAS+CADD_PER | 0.7184 | 0.6950 | 0.4000 | 0.6900 | 0.5800 | 0.7436 | 0.6517 | 0.3895 |
| Baseline | 0.5680 | 0.5951 | 0.1600 | 0.5800 | 0.4800 | 0.6000 | 0.5330 | 0.1633 |
| Baseline+GWAS | 0.6592 | 0.6634 | 0.3200 | 0.6600 | 0.4400 | 0.7857 | 0.5641 | 0.3563 |

**Supplementary Table 4.** Performance metrics (AUROC, AUPR, KS, ACC, recall, precision, F1, MCC) for ELAG and baseline methods across different patterns (Baseline, Basline+GWAS, ELAG, ELAG+GWAS, ELAG+GWAS+CADD, ELAG+GWAS+CADD_PER), with a heritability of 0.2 and a sample size of 1,000. CADD_PER means CADD scores are permutated. Simulations included 1,000 causal QTLs, with half exhibiting linear effects and half nonlinear effects.

| PRS Methods | Feature Selection | AUROC | AUPR | KS | ACC | Recall | Precision | F1 Score | MCC |
|---|---|---|---|---|---|---|---|---|---|
| PRSice | ELAG | 0.8192 | 0.8180 | 0.5400 | 0.7700 | 0.7000 | 0.8140 | 0.7527 | 0.5454 |
| | Original | 0.7640 | 0.7700 | 0.4200 | 0.7100 | 0.6200 | 0.7561 | 0.6813 | 0.4270 |
| Lassosum | ELAG | 0.7492 | 0.7312 | 0.5200 | 0.7600 | 0.7800 | 0.7500 | 0.7647 | 0.5204 |
| | Original | 0.6864 | 0.7013 | 0.3200 | 0.6600 | 0.4000 | 0.8333 | 0.5405 | 0.3746 |

**Supplementary Table 5.** Performance metrics (AUROC, AUPR, KS, ACC, recall, precision, F1, MCC) for ELAG+PRS and original PRS methods (PRSice and Lassosum), with a heritability of 0.2 and a sample size of 1,000. Simulations included 1,000 causal QTLs, with half exhibiting linear effects and half nonlinear effects.

| Sample Size | Method | AUROC | AUPR | KS | ACC | MCC |
|---|---|---|---|---|---|---|
| 1000 | ELAG | 0.6324 | 0.6405 | 0.28 | 0.63 | 0.26926 |
| | MWPG | 0.6008 | 0.6182 | 0.2 | 0.59 | 0.2209 |
| | Baseline | 0.568 | 0.5950726 | 0.16 | 0.58 | 0.1633 |
| 3000 | ELAG | 0.67036 | 0.6432 | 0.2933 | 0.64333 | 0.2872 |
| | MWPG | 0.6563 | 0.6479 | 0.247 | 0.613 | 0.228 |
| | Baseline | 0.6308 | 0.6362 | 0.2133 | 0.6067 | 0.2143 |
| 10000 | ELAG | 0.718 | 0.6863 | 0.35 | 0.673 | 0.3481 |
| | MWPG | 0.7123 | 0.6869 | 0.322 | 0.658 | 0.322 |
| | Baseline | 0.692732 | 0.6629 | 0.292 | 0.643 | 0.2954 |

**Supplementary Table 6.** Main performance metrics in the ablation study comparing ELAG, MWPG, and the baseline method. Simulations are with $h^2$=0.2 and 1,000 causal QTLs.

| Disease | Feature Selection | AUROC | AUPR | KS | ACC | Recall | Precision | F1 Score | MCC |
|---|---|---|---|---|---|---|---|---|---|
| Alzheimer's Disease | ELAG+GWAS | 0.7237 | 0.7421 | 0.3521 | 0.6714 | 0.7793 | 0.641 | 0.7034 | 0.351 |
| | ELAG+GWAS +CADD | 0.7199 | 0.745 | 0.3474 | 0.6737 | 0.7277 | 0.6568 | 0.6904 | 0.3495 |
| Rheumatoid Arthritis | ELAG+GWAS | 0.7354 | 0.7284 | 0.3663 | 0.6832 | 0.8218 | 0.6434 | 0.7217 | 0.3813 |
| | ELAG+GWAS +CADD | 0.7214 | 0.701 | 0.3762 | 0.6831 | 0.7723 | 0.6555 | 0.7091 | 0.3723 |
| Multiple Sclerosis | ELAG+GWAS | 0.7185 | 0.6953 | 0.3537 | 0.6768 | 0.7256 | 0.6611 | 0.6919 | 0.3533 |
| | ELAG+GWAS +CADD | 0.7104 | 0.7022 | 0.3293 | 0.6646 | 0.6768 | 0.6607 | 0.6687 | 0.3294 |

**Supplementary Table 7.** Performance metrics (AUROC, AUPR, KS, ACC, recall, precision, F1, MCC) for ELAG with GWAS statistics and GWAS statistic + CADD.

| Disease | Missing Rate | AUROC | AUPR | KS | ACC | Recall | Precision | F1 Score | MCC |
|---|---|---|---|---|---|---|---|---|---|
| Alzheimer's Disease | 20% | 0.7169 | 0.7302 | 0.3474 | 0.6714 | 0.6714 | 0.6714 | 0.6714 | 0.3427 |
| | 40% | 0.7068 | 0.7293 | 0.3333 | 0.6643 | 0.6244 | 0.6786 | 0.6504 | 0.3297 |
| | 60% | 0.7052 | 0.7287 | 0.3474 | 0.6714 | 0.6479 | 0.6798 | 0.6635 | 0.3431 |
| | 80% | 0.6983 | 0.7220 | 0.3474 | 0.6620 | 0.5117 | 0.7315 | 0.6022 | 0.3396 |
| Rheumatoid Arthritis | 20% | 0.7346 | 0.7184 | 0.3762 | 0.6832 | 0.9307 | 0.6225 | 0.7460 | 0.4216 |
| | 40% | 0.7292 | 0.7295 | 0.3465 | 0.6683 | 0.8911 | 0.6164 | 0.7287 | 0.3760 |
| | 60% | 0.7284 | 0.7235 | 0.3663 | 0.6733 | 0.6436 | 0.6842 | 0.6633 | 0.3471 |
| | 80% | 0.7069 | 0.7178 | 0.3267 | 0.6584 | 0.7723 | 0.6290 | 0.6933 | 0.3254 |
| Multiple Sclerosis | 20% | 0.7170 | 0.6945 | 0.3598 | 0.6768 | 0.6098 | 0.7042 | 0.6536 | 0.3569 |
| | 40% | 0.7124 | 0.6875 | 0.3476 | 0.6646 | 0.7927 | 0.6311 | 0.7027 | 0.3406 |
| | 60% | 0.7013 | 0.6831 | 0.3171 | 0.6585 | 0.5549 | 0.7000 | 0.6190 | 0.3241 |
| | 80% | 0.6879 | 0.6600 | 0.2805 | 0.6280 | 0.7988 | 0.5955 | 0.6823 | 0.2725 |

**Supplementary Table 8.** Performance metrics (AUROC, AUPR, KS, ACC, recall, precision, F1, MCC) of ELAG under varying levels of individual genotype missingness (20%, 40%, 60%, 80%) across three diseases.

| Disease | Features | AUROC | AUPR | KS | ACC | Recall | Precision | F1_Score | MCC |
|---|---|---|---|---|---|---|---|---|---|
| AD | All features | 0.8658 | 0.8584 | 0.5962 | 0.7958 | 0.8216 | 0.7813 | 0.8009 | 0.5923 |
| | Only age/sex | 0.8183 | 0.7778 | 0.4883 | 0.7441 | 0.7793 | 0.7281 | 0.7528 | 0.4895 |
| | Only genetics | 0.7238 | 0.7421 | 0.3521 | 0.6714 | 0.7793 | 0.6409 | 0.7034 | 0.3510 |
| RA | All features | 0.7420 | 0.7523 | 0.3366 | 0.6683 | 0.8020 | 0.6328 | 0.7074 | 0.3493 |
| | Only age/sex | 0.6598 | 0.6477 | 0.3069 | 0.6535 | 0.6733 | 0.6476 | 0.6602 | 0.3072 |
| | Only genetics | 0.7354 | 0.7284 | 0.3663 | 0.6832 | 0.8218 | 0.6434 | 0.7217 | 0.3813 |
| MS | All features | 0.7363 | 0.7243 | 0.3902 | 0.6921 | 0.6037 | 0.7333 | 0.6622 | 0.3903 |
| | Only age/sex | 0.5460 | 0.5427 | 0.0976 | 0.5488 | 0.7988 | 0.5325 | 0.6390 | 0.1127 |
| | Only genetics | 0.7185 | 0.6953 | 0.3537 | 0.6768 | 0.7256 | 0.6611 | 0.6919 | 0.3533 |

**Supplementary Table 15.** Performance metrics of the ELAG models across feature sets and diseases. The table displays eight evaluation metrics for Alzheimer's Disease (AD), Rheumatoid Arthritis (RA), and Multiple Sclerosis (MS) using three feature configurations.
